## supplemental tables and figures for "The effect of strict lockdown on Omicron SARS-CoV-2 variant transmission in Shanghai"

Supplementary Table 1. Calculating sub-index scores for indicators of metrics.

| **Name** | **Coding** | **Score** |
| --- | --- | --- |
| **C1(School closing)** | 0 - no measures | 0 |
|  | 1 - recommend closing or all schools open with alterations resulting in significant differences compared to non-Covid-19 operations | 33.33 |
|  | 2 - require closing (only some levels or categories, eg just high school, or just public schools) | 66.67 |
|  | 3 - require closing all levels | 100 |
| **C2(Workplace closing)** | 0 - no measures | 0 |
|  | 1 - recommend closing (or recommend work from home) or all businesses open with alterations resulting in significant differences compared to non-Covid-19 operation | 33.33 |
|  | 2 - require closing (or work from home) for some sectors or categories of workers | 66.67 |
|  | 3 - require closing (or work from home) for all-but-essential workplaces (eg grocery stores, doctors) | 100 |
| **C3(Cancel public events)** | 0 - no measures | 0 |
|  | 1 - recommend cancelling | 50 |
|  | 2 - require cancelling | 100 |
| **C4(Restrictions on gatherings)** | 0 - no restrictions | 0 |
|  | 1 - restrictions on very large gatherings (the limit is above 1000 people) | 25 |
|  | 2 - restrictions on gatherings between 101-1000 people | 50 |
|  | 3 - restrictions on gatherings between 11-100 people | 75 |
|  | 4 - restrictions on gatherings of 10 people or less | 100 |
| **C5(Close public transport)** | 0 - no measures | 0 |
|  | 1 - recommend closing (or significantly reduce volume/route/means of transport available) | 50 |
|  | 2 - require closing (or prohibit most citizens from using it) | 100 |
| **C6(Stay at home requirements)** | 0 - no measures | 0 |
|  | 1 - recommend not leaving house | 33.33 |
|  | 2 - require not leaving house with exceptions for daily exercise, grocery shopping, and 'essential' trips | 66.67 |
|  | 3 - require not leaving house with minimal exceptions (eg allowed to leave once a week, or only one person can leave at a time, etc) | 100 |
| **C7(Restrictions on internal movement)** | 0 - no measures | 0 |
|  | 1 - recommend not to travel between regions/cities | 50 |
|  | 2 - internal movement restrictions in place | 100 |
| **C8(International travel controls)** | 0 - no restrictions | 0 |
|  | 1 - screening arrivals | 25 |
|  | 2 - quarantine arrivals from some or all regions | 50 |
|  | 3 - ban arrivals from some regions | 75 |
|  | 4 - ban on all regions or total border closure | 100 |

The scores of each indicator are calculated by the equation described in the website link (https://github.com/OxCGRT/covid-policy-tracker/blob/master/documentation/index_methodology. md).

Supplementary Table 2. Stringency indices of lockdown policies implemented by different countries.

| **Country** | **C1** | | **C2** | | **C3** | | **C4** | | **C5** | | **C6** | | **C7** | | **C8** | | **Stringency indices** | |
| --- | --- | --- | --- | --- | --- | --- | --- | --- | --- | --- | --- | --- | --- | --- | --- | --- | --- | --- |
| **India** | | 100 | | 100 | | 100 | | 100 | | 100 | | 100 | | 100 | | 100 | | 100 |
| **Italy** | | 100 | | 100 | | 100 | | 100 | | 100 | | 100 | | 100 | | 75 | | 96.88 |
| **Shanghai, China** | | 100 | | 100 | | 100 | | 100 | | 100 | | 100 | | 100 | | 75 | | 96.88 |
| **Germany** | | 100 | | 100 | | 100 | | 100 | | 50 | | 100 | | 100 | | 75 | | 90.62 |
| **France** | | 100 | | 100 | | 100 | | 100 | | 50 | | 100 | | 100 | | 75 | | 90.62 |
| **United States** | | 100 | | 100 | | 100 | | 100 | | 50 | | 100 | | 100 | | 75 | | 90.62 |
| **Australia** | | 100 | | 66.67 | | 100 | | 100 | | 50 | | 100 | | 100 | | 100 | | 89.58 |
| **Canada** | | 100 | | 100 | | 100 | | 100 | | 0 | | 100 | | 100 | | 100 | | 87.5 |
| **Spain** | | 100 | | 100 | | 100 | | 100 | | 50 | | 100 | | 50 | | 100 | | 87.5 |
| **Netherland** | | 100 | | 100 | | 100 | | 100 | | 50 | | 100 | | 50 | | 75 | | 84.37 |
| **United Kingdom** | | 100 | | 100 | | 100 | | 100 | | 50 | | 100 | | 100 | | 0 | | 81.25 |
| **Singapore** | | 100 | | 100 | | 100 | | 100 | | 0 | | 66.67 | | 100 | | 75 | | 80.21 |
| **South Korea** | | 100 | | 100 | | 100 | | 100 | | 0 | | 66.67 | | 100 | | 75 | | 80.21 |
| **Norway** | | 100 | | 66.67 | | 100 | | 100 | | 50 | | 0 | | 100 | | 100 | | 77.08 |
| **Finland** | | 66.67 | | 66.67 | | 100 | | 100 | | 0 | | 33.33 | | 100 | | 100 | | 70.83 |
| **Denmark** | | 100 | | 66.67 | | 50 | | 100 | | 50 | | 33.33 | | 50 | | 100 | | 68.75 |
| **Sweden** | | 66.67 | | 66.67 | | 100 | | 100 | | 50 | | 33.33 | | 50 | | 75 | | 67.71 |
| **Japan** | | 66.67 | | 66.67 | | 100 | | 50 | | 0 | | 33.33 | | 50 | | 100 | | 58.33 |

Stringency indices are simple averages of the individual component indicators.

Supplementary Table 3. Daily new infections in Shanghai from March 1^st^ to April 30^th^, 2022.

| **Date** | **Daily symptomatic cases** | **Daily asymptomatic cases** | **Daily new infections** |
| --- | --- | --- | --- |
| **Mar 01, 2022** | 1 | 18 | 19 |
| **Mar 02, 2022** | 3 | 24 | 27 |
| **Mar 03, 2022** | 2 | 35 | 37 |
| **Mar 04, 2022** | 3 | 26 | 29 |
| **Mar 05, 2022** | 0 | 38 | 38 |
| **Mar 06, 2022** | 3 | 61 | 64 |
| **Mar 07, 2022** | 4 | 61 | 65 |
| **Mar 08, 2022** | 3 | 72 | 75 |
| **Mar 09, 2022** | 4 | 92 | 96 |
| **Mar 10, 2022** | 11 | 74 | 85 |
| **Mar 11, 2022** | 5 | 87 | 92 |
| **Mar 12, 2022** | 1 | 65 | 66 |
| **Mar 13, 2022** | 41 | 130 | 171 |
| **Mar 14, 2022** | 9 | 133 | 142 |
| **Mar 15, 2022** | 5 | 198 | 203 |
| **Mar 16, 2022** | 8 | 156 | 164 |
| **Mar 17, 2022** | 57 | 205 | 262 |
| **Mar 18, 2022** | 8 | 372 | 380 |
| **Mar 19, 2022** | 17 | 494 | 511 |
| **Mar 20, 2022** | 24 | 736 | 760 |
| **Mar 21, 2022** | 31 | 868 | 899 |
| **Mar 22, 2022** | 4 | 981 | 985 |
| **Mar 23, 2022** | 4 | 982 | 986 |
| **Mar 24, 2022** | 29 | 1587 | 1616 |
| **Mar 25, 2022** | 38 | 2233 | 2271 |
| **Mar 26, 2022** | 45 | 2633 | 2678 |
| **Mar 27, 2022** | 50 | 3454 | 3504 |
| **Mar 28, 2022** | 96 | 4382 | 4478 |
| **Mar 29, 2022** | 326 | 5658 | 5984 |
| **Mar 30, 2022** | 355 | 5299 | 5654 |
| **Mar 31, 2022** | 358 | 4145 | 4503 |
| **Apr 01, 2022** | 260 | 6051 | 6311 |
| **Apr 02, 2022** | 438 | 7789 | 8227 |
| **Apr 03, 2022** | 425 | 8585 | 9010 |
| **Apr 04, 2022** | 268 | 13088 | 13356 |
| **Apr 05, 2022** | 311 | 16767 | 17078 |
| **Apr 06, 2022** | 322 | 19661 | 19983 |
| **Apr 07, 2022** | 824 | 20401 | 21225 |
| **Apr 08, 2022** | 1015 | 22611 | 23626 |
| **Apr 09, 2022** | 1006 | 23939 | 24945 |
| **Apr 10, 2022** | 914 | 25173 | 26087 |
| **Apr 11, 2022** | 994 | 22349 | 23343 |
| **Apr 12, 2022** | 1189 | 25141 | 26330 |
| **Apr 13, 2022** | 2573 | 25147 | 27720 |
| **Apr 14, 2022** | 3200 | 19873 | 23073 |
| **Apr 15, 2022** | 3590 | 19925 | 23515 |
| **Apr 16, 2022** | 3238 | 21582 | 24820 |
| **Apr 17, 2022** | 2417 | 19831 | 22248 |
| **Apr 18, 2022** | 3084 | 17334 | 20418 |
| **Apr 19, 2022** | 2494 | 16408 | 18902 |
| **Apr 20, 2022** | 2634 | 15863 | 18497 |
| **Apr 21, 2022** | 1931 | 15698 | 17629 |
| **Apr 22, 2022** | 2736 | 20635 | 23371 |
| **Apr 23, 2022** | 1401 | 19657 | 21058 |
| **Apr 24, 2022** | 2472 | 16984 | 19456 |
| **Apr 25, 2022** | 1661 | 15319 | 16980 |
| **Apr 26, 2022** | 1606 | 11956 | 13562 |
| **Apr 27, 2022** | 1292 | 9330 | 10622 |
| **Apr 28, 2022** | 5487 | 9547 | 15034 |
| **Apr 29, 2022** | 1249 | 8932 | 10181 |
| **Apr 30, 2022** | 788 | 7086 | 7874 |

Daily new infections are the sum of symptomatic and asymptomatic cases.

Supplementary Table 4. Daily stringency index in Shanghai.

| **Date** | **C1** | **C2** | **C3** | **C4** | **C5** | **C6** | **C7** | **C8** | **Stringency index** |
| --- | --- | --- | --- | --- | --- | --- | --- | --- | --- |
| **Mar 01, 2022** | 0 | 0 | 0 | 0 | 0 | 0 | 0 | 75 | 9.38 |
| **Mar 02, 2022** | 0 | 0 | 0 | 0 | 0 | 0 | 0 | 75 | 9.38 |
| **Mar 03, 2022** | 0 | 0 | 0 | 0 | 0 | 0 | 0 | 75 | 9.38 |
| **Mar 04, 2022** | 0 | 0 | 0 | 0 | 0 | 0 | 0 | 75 | 9.38 |
| **Mar 05, 2022** | 0 | 0 | 0 | 0 | 0 | 0 | 0 | 75 | 9.38 |
| **Mar 06, 2022** | 0 | 0 | 0 | 0 | 0 | 0 | 0 | 75 | 9.38 |
| **Mar 07, 2022** | 0 | 0 | 0 | 0 | 0 | 0 | 0 | 75 | 9.38 |
| **Mar 08, 2022** | 0 | 0 | 0 | 0 | 0 | 0 | 0 | 75 | 9.38 |
| **Mar 09, 2022** | 0 | 0 | 0 | 0 | 0 | 0 | 0 | 75 | 9.38 |
| **Mar 10, 2022** | 0 | 0 | 0 | 0 | 0 | 0 | 0 | 75 | 9.38 |
| **Mar 11, 2022** | 0 | 0 | 0 | 0 | 0 | 0 | 0 | 75 | 9.38 |
| **Mar 12, 2023** | 0 | 0 | 0 | 0 | 0 | 0 | 0 | 75 | 9.38 |
| **Mar 13, 2022** | 66.67 | 33.33 | 50 | 50 | 50 | 33.33 | 0 | 75 | 44.79 |
| **Mar 14, 2022** | 66.67 | 33.33 | 50 | 50 | 50 | 33.33 | 0 | 75 | 44.79 |
| **Mar 15, 2022** | 66.67 | 33.33 | 50 | 50 | 50 | 33.33 | 0 | 75 | 44.79 |
| **Mar 16, 2022** | 66.67 | 33.33 | 50 | 50 | 50 | 33.33 | 0 | 75 | 44.79 |
| **Mar 17, 2022** | 66.67 | 33.33 | 50 | 50 | 50 | 33.33 | 0 | 75 | 44.79 |
| **Mar 18, 2022** | 66.67 | 33.33 | 50 | 50 | 50 | 33.33 | 0 | 75 | 44.79 |
| **Mar 19, 2022** | 66.67 | 33.33 | 50 | 50 | 50 | 33.33 | 0 | 75 | 44.79 |
| **Mar 20, 2022** | 66.67 | 33.33 | 50 | 50 | 50 | 33.33 | 0 | 75 | 44.79 |
| **Mar 21, 2022** | 66.67 | 33.33 | 50 | 50 | 50 | 33.33 | 0 | 75 | 44.79 |
| **Mar 22, 2022** | 66.67 | 33.33 | 50 | 50 | 50 | 33.33 | 0 | 75 | 44.79 |
| **Mar 23, 2022** | 66.67 | 33.33 | 50 | 50 | 50 | 33.33 | 0 | 75 | 44.79 |
| **Mar 24, 2022** | 66.67 | 33.33 | 50 | 50 | 50 | 33.33 | 0 | 75 | 44.79 |
| **Mar 25, 2022** | 66.67 | 33.33 | 50 | 50 | 50 | 33.33 | 0 | 75 | 44.79 |
| **Mar 26, 2022** | 66.67 | 33.33 | 50 | 50 | 50 | 33.33 | 0 | 75 | 44.79 |
| **Mar 27, 2022** | 66.67 | 33.33 | 50 | 50 | 50 | 33.33 | 0 | 75 | 44.79 |
| **Mar 28, 2022** | 83.33 | 50 | 75 | 62.50 | 75 | 83.33 | 25 | 75 | 66.15 |
| **Mar 29, 2022** | 83.33 | 50 | 75 | 62.50 | 75 | 83.33 | 25 | 75 | 66.15 |
| **Mar 30, 2022** | 83.33 | 50 | 75 | 62.50 | 75 | 83.33 | 25 | 75 | 66.15 |
| **Mar 31, 2022** | 83.33 | 50 | 75 | 62.50 | 75 | 83.33 | 25 | 75 | 66.15 |
| **Apr 01, 2022** | 100 | 100 | 100 | 100 | 100 | 100 | 100 | 75 | 96.88 |
| **Apr 02, 2022** | 100 | 100 | 100 | 100 | 100 | 100 | 100 | 75 | 96.88 |
| **Apr 03, 2022** | 100 | 100 | 100 | 100 | 100 | 100 | 100 | 75 | 96.88 |
| **Apr 04, 2022** | 100 | 100 | 100 | 100 | 100 | 100 | 100 | 75 | 96.88 |
| **Apr 05, 2022** | 100 | 100 | 100 | 100 | 100 | 100 | 100 | 75 | 96.88 |
| **Apr 06, 2022** | 100 | 100 | 100 | 100 | 100 | 100 | 100 | 75 | 96.88 |
| **Apr 07, 2022** | 100 | 100 | 100 | 100 | 100 | 100 | 100 | 75 | 96.88 |
| **Apr 08, 2022** | 100 | 100 | 100 | 100 | 100 | 100 | 100 | 75 | 96.88 |
| **Apr 09, 2022** | 100 | 100 | 100 | 100 | 100 | 100 | 100 | 75 | 96.88 |
| **Apr 10, 2022** | 100 | 100 | 100 | 100 | 100 | 100 | 100 | 75 | 96.88 |
| **Apr 11, 2022** | 100 | 100 | 100 | 100 | 100 | 100 | 100 | 75 | 96.88 |
| **Apr 12, 2022** | 100 | 100 | 100 | 100 | 100 | 100 | 100 | 75 | 96.88 |
| **Apr 13, 2022** | 100 | 100 | 100 | 100 | 100 | 100 | 100 | 75 | 96.88 |
| **Apr 14, 2022** | 100 | 100 | 100 | 100 | 100 | 100 | 100 | 75 | 96.88 |
| **Apr 15, 2022** | 100 | 100 | 100 | 100 | 100 | 100 | 100 | 75 | 96.88 |
| **Apr 16, 2022** | 100 | 100 | 100 | 100 | 100 | 100 | 100 | 75 | 96.88 |
| **Apr 17, 2022** | 100 | 100 | 100 | 100 | 100 | 100 | 100 | 75 | 96.88 |
| **Apr 18, 2022** | 100 | 100 | 100 | 100 | 100 | 100 | 100 | 75 | 96.88 |
| **Apr 19, 2022** | 100 | 100 | 100 | 100 | 100 | 100 | 100 | 75 | 96.88 |
| **Apr 20, 2022** | 100 | 100 | 100 | 100 | 100 | 100 | 100 | 75 | 96.88 |
| **Apr 21, 2022** | 100 | 100 | 100 | 100 | 100 | 100 | 100 | 75 | 96.88 |
| **Apr 22, 2022** | 100 | 100 | 100 | 100 | 100 | 100 | 100 | 75 | 96.88 |
| **Apr 23, 2022** | 100 | 100 | 100 | 100 | 100 | 100 | 100 | 75 | 96.88 |
| **Apr 24, 2022** | 100 | 100 | 100 | 100 | 100 | 100 | 100 | 75 | 96.88 |
| **Apr 25, 2022** | 100 | 100 | 100 | 100 | 100 | 100 | 100 | 75 | 96.88 |
| **Apr 26, 2022** | 100 | 100 | 100 | 100 | 100 | 100 | 100 | 75 | 96.88 |
| **Apr 27, 2022** | 100 | 100 | 100 | 100 | 100 | 100 | 100 | 75 | 96.88 |
| **Apr 28, 2022** | 100 | 100 | 100 | 100 | 100 | 100 | 100 | 75 | 96.88 |
| **Apr 29, 2022** | 100 | 100 | 100 | 100 | 100 | 100 | 100 | 75 | 96.88 |
| **Apr 30, 2022** | 100 | 100 | 100 | 100 | 100 | 100 | 100 | 75 | 96.88 |

Supplementary Table 5. Daily new infections in the 16 districts of Shanghai.

| **Date** | **Jingan** | **Huangpu** | **Pudong** | **Hongkou** | **Changning** | **Putuo** | **Xuhui** | **Yangpu** |
| --- | --- | --- | --- | --- | --- | --- | --- | --- |
| **Mar 01** | 0 | 0 | 0 | 0 | 0 | 1 | 0 | 0 |
| **Mar 02** | 0 | 0 | 0 | 0 | 0 | 6 | 0 | 0 |
| **Mar 03** | 0 | 0 | 0 | 0 | 0 | 9 | 0 | 0 |
| **Mar 04** | 0 | 1 | 0 | 0 | 0 | 1 | 0 | 0 |
| **Mar 05** | 0 | 1 | 2 | 0 | 0 | 1 | 3 | 0 |
| **Mar 06** | 1 | 1 | 2 | 0 | 0 | 5 | 5 | 0 |
| **Mar 07** | 0 | 0 | 2 | 0 | 0 | 0 | 20 | 1 |
| **Mar 08** | 2 | 0 | 3 | 0 | 1 | 2 | 9 | 0 |
| **Mar 09** | 5 | 2 | 10 | 1 | 1 | 1 | 7 | 2 |
| **Mar 10** | 2 | 1 | 10 | 6 | 1 | 4 | 8 | 0 |
| **Mar 11** | 3 | 3 | 9 | 3 | 1 | 0 | 12 | 0 |
| **Mar 12** | 1 | 3 | 35 | 0 | 2 | 1 | 1 | 0 |
| **Mar 13** | 14 | 13 | 37 | 4 | 1 | 7 | 18 | 3 |
| **Mar 14** | 0 | 2 | 1 | 1 | 0 | 0 | 3 | 0 |
| **Mar 15** | 0 | 0 | 2 | 0 | 0 | 0 | 2 | 0 |
| **Mar 16** | 3 | 9 | 43 | 10 | 4 | 4 | 7 | 2 |
| **Mar 17** | 5 | 23 | 35 | 6 | 1 | 11 | 35 | 3 |
| **Mar 18** | 13 | 15 | 65 | 19 | 7 | 22 | 41 | 8 |
| **Mar 19** | 15 | 37 | 135 | 7 | 12 | 25 | 60 | 14 |
| **Mar 20** | 0 | 2 | 1 | 0 | 1 | 0 | 1 | 0 |
| **Mar 21** | 45 | 48 | 170 | 23 | 26 | 32 | 133 | 10 |
| **Mar 22** | 28 | 59 | 237 | 13 | 20 | 21 | 86 | 3 |
| **Mar 23** | 48 | 20 | 217 | 26 | 11 | 34 | 108 | 11 |
| **Mar 24** | 71 | 102 | 195 | 29 | 59 | 38 | 167 | 21 |
| **Mar 25** | 2 | 23 | 1915 | 4 | 4 | 9 | 3 | 11 |
| **Mar 26** | 108 | 159 | 323 | 49 | 29 | 69 | 332 | 24 |
| **Mar 27** | 70 | 56 | 1424 | 27 | 46 | 71 | 280 | 35 |
| **Mar 28** | 103 | 283 | 2502 | 8 | 115 | 82 | 92 | 49 |
| **Mar 29** | 180 | 112 | 2187 | 73 | 36 | 112 | 1088 | 98 |
| **Mar 30** | 100 | 363 | 2198 | 84 | 130 | 148 | 400 | 99 |
| **Mar 31** | 162 | 124 | 2404 | 126 | 258 | 17 | 226 | 174 |
| **Apr 01** | 189 | 247 | 2597 | 60 | 38 | 249 | 631 | 135 |
| **Apr 02** | 326 | 658 | 2056 | 344 | 100 | 389 | 1035 | 275 |
| **Apr 03** | 344 | 827 | 3669 | 189 | 104 | 325 | 498 | 374 |
| **Apr 04** | 60 | 981 | 6993 | 609 | 39 | 259 | 1231 | 225 |
| **Apr 05** | 317 | 663 | 8054 | 411 | 92 | 489 | 925 | 626 |
| **Apr 06** | 543 | 1050 | 8447 | 670 | 359 | 1035 | 1106 | 633 |
| **Apr 07** | 382 | 1382 | 9041 | 594 | 854 | 958 | 2073 | 601 |
| **Apr 08** | 665 | 2607 | 7286 | 378 | 614 | 1094 | 1629 | 701 |
| **Apr 09** | 602 | 548 | 11130 | 349 | 757 | 653 | 1150 | 1080 |
| **Apr 10** | 603 | 1761 | 6732 | 1238 | 405 | 1001 | 3185 | 1877 |
| **Apr 11** | 552 | 2223 | 8306 | 1375 | 393 | 1878 | 1771 | 1420 |
| **Apr 12** | 1056 | 1804 | 11049 | 931 | 1133 | 1170 | 1108 | 1159 |
| **Apr 13** | 220 | 1408 | 15027 | 1488 | 957 | 475 | 1492 | 1185 |
| **Apr 14** | 266 | 2013 | 11656 | 930 | 998 | 264 | 1342 | 817 |
| **Apr 15** | 433 | 1384 | 10282 | 1497 | 752 | 426 | 1689 | 1411 |
| **Apr 16** | 1134 | 1565 | 10791 | 1027 | 706 | 1331 | 1532 | 860 |
| **Apr 17** | 842 | 1896 | 7740 | 1166 | 739 | 1225 | 1553 | 887 |
| **Apr 18** | 1010 | 3323 | 8831 | 1092 | 680 | 371 | 572 | 481 |
| **Apr 19** | 1733 | 3084 | 5646 | 753 | 618 | 528 | 1608 | 766 |
| **Apr 20** | 1130 | 3097 | 4465 | 832 | 726 | 555 | 1447 | 2005 |
| **Apr 21** | 2064 | 1752 | 4655 | 885 | 496 | 489 | 1366 | 543 |
| **Apr 22** | 786 | 2843 | 7961 | 997 | 713 | 523 | 1275 | 1356 |
| **Apr 23** | 471 | 1959 | 7626 | 819 | 727 | 478 | 1722 | 1277 |
| **Apr 24** | 1702 | 1755 | 6181 | 655 | 749 | 477 | 1396 | 1235 |
| **Apr 25** | 1016 | 2671 | 3912 | 1270 | 534 | 418 | 1341 | 1244 |
| **Apr 26** | 1394 | 1228 | 2745 | 1136 | 419 | 365 | 904 | 1106 |
| **Apr 27** | 1089 | 1452 | 2993 | 1091 | 208 | 301 | 552 | 526 |
| **Apr 28** | 1364 | 2656 | 3472 | 1374 | 647 | 235 | 1580 | 808 |
| **Apr 29** | 1267 | 1399 | 2028 | 925 | 377 | 141 | 602 | 767 |
| **Apr 30** | 996 | 1156 | 1625 | 591 | 181 | 114 | 488 | 342 |

| **Date** | **Chongming** | **Minhang** | **Jinshan** | **Baoshan** | **Songjiang** | **Qingpu** | **Jiading** | **Fengxian** |
| --- | --- | --- | --- | --- | --- | --- | --- | --- |
| **Mar 01** | 0 | 0 | 0 | 0 | 0 | 0 | 1 | 0 |
| **Mar 02** | 0 | 0 | 0 | 1 | 0 | 0 | 1 | 0 |
| **Mar 03** | 0 | 0 | 0 | 2 | 5 | 0 | 0 | 0 |
| **Mar 04** | 0 | 1 | 0 | 1 | 6 | 2 | 7 | 0 |
| **Mar 05** | 0 | 6 | 0 | 2 | 7 | 1 | 4 | 1 |
| **Mar 06** | 0 | 7 | 0 | 6 | 7 | 0 | 12 | 2 |
| **Mar 07** | 0 | 10 | 1 | 5 | 3 | 0 | 13 | 0 |
| **Mar 08** | 0 | 12 | 0 | 11 | 14 | 3 | 8 | 0 |
| **Mar 09** | 0 | 23 | 0 | 2 | 9 | 0 | 16 | 1 |
| **Mar 10** | 0 | 15 | 0 | 11 | 9 | 4 | 4 | 0 |
| **Mar 11** | 0 | 20 | 4 | 8 | 5 | 2 | 12 | 1 |
| **Mar 12** | 0 | 11 | 4 | 3 | 1 | 0 | 3 | 0 |
| **Mar 13** | 0 | 28 | 7 | 4 | 7 | 4 | 20 | 2 |
| **Mar 14** | 0 | 1 | 0 | 0 | 1 | 0 | 0 | 0 |
| **Mar 15** | 0 | 1 | 0 | 0 | 0 | 0 | 0 | 0 |
| **Mar 16** | 0 | 18 | 0 | 11 | 4 | 1 | 40 | 2 |
| **Mar 17** | 3 | 79 | 1 | 10 | 10 | 1 | 34 | 3 |
| **Mar 18** | 0 | 60 | 17 | 27 | 13 | 16 | 44 | 7 |
| **Mar 19** | 23 | 53 | 3 | 17 | 16 | 5 | 81 | 6 |
| **Mar 20** | 0 | 18 | 0 | 0 | 0 | 0 | 1 | 0 |
| **Mar 21** | 40 | 120 | 16 | 66 | 38 | 5 | 102 | 22 |
| **Mar 22** | 19 | 306 | 3 | 16 | 31 | 20 | 111 | 8 |
| **Mar 23** | 54 | 254 | 5 | 69 | 34 | 10 | 69 | 13 |
| **Mar 24** | 82 | 489 | 12 | 87 | 92 | 13 | 130 | 24 |
| **Mar 25** | 41 | 206 | 1 | 4 | 15 | 13 | 18 | 0 |
| **Mar 26** | 27 | 970 | 4 | 153 | 82 | 20 | 242 | 85 |
| **Mar 27** | 237 | 616 | 14 | 94 | 190 | 43 | 252 | 45 |
| **Mar 28** | 68 | 369 | 19 | 312 | 95 | 56 | 209 | 115 |
| **Mar 29** | 56 | 986 | 26 | 362 | 234 | 84 | 253 | 95 |
| **Mar 30** | 195 | 780 | 33 | 507 | 248 | 78 | 160 | 130 |
| **Mar 31** | 43 | 389 | 43 | 17 | 190 | 98 | 54 | 177 |
| **Apr 01** | 84 | 1029 | 50 | 46 | 507 | 87 | 200 | 162 |
| **Apr 02** | 111 | 822 | 61 | 480 | 573 | 224 | 614 | 158 |
| **Apr 03** | 144 | 937 | 103 | 463 | 264 | 220 | 424 | 121 |
| **Apr 04** | 49 | 1381 | 53 | 268 | 569 | 320 | 247 | 70 |
| **Apr 05** | 78 | 2940 | 82 | 555 | 801 | 400 | 491 | 153 |
| **Apr 06** | 63 | 2408 | 79 | 662 | 783 | 469 | 1408 | 267 |
| **Apr 07** | 262 | 2255 | 128 | 416 | 754 | 495 | 934 | 93 |
| **Apr 08** | 170 | 2854 | 90 | 2821 | 771 | 342 | 1505 | 97 |
| **Apr 09** | 171 | 4624 | 68 | 2261 | 500 | 557 | 369 | 124 |
| **Apr 10** | 55 | 3189 | 57 | 1839 | 1825 | 877 | 1405 | 38 |
| **Apr 11** | 57 | 3007 | 38 | 1024 | 691 | 331 | 208 | 68 |
| **Apr 12** | 86 | 4245 | 39 | 295 | 712 | 524 | 986 | 33 |
| **Apr 13** | 63 | 2939 | 34 | 651 | 657 | 319 | 721 | 83 |
| **Apr 14** | 60 | 2378 | 28 | 417 | 761 | 304 | 803 | 35 |
| **Apr 15** | 92 | 2037 | 31 | 1323 | 773 | 536 | 741 | 106 |
| **Apr 16** | 49 | 3060 | 21 | 1293 | 410 | 305 | 685 | 51 |
| **Apr 17** | 52 | 2402 | 24 | 1450 | 905 | 369 | 978 | 20 |
| **Apr 18** | 26 | 1372 | 7 | 1030 | 762 | 257 | 588 | 14 |
| **Apr 19** | 80 | 1602 | 9 | 957 | 325 | 389 | 743 | 60 |
| **Apr 20** | 65 | 1686 | 25 | 833 | 676 | 280 | 641 | 32 |
| **Apr 21** | 120 | 2563 | 30 | 978 | 493 | 385 | 776 | 34 |
| **Apr 22** | 117 | 1305 | 26 | 2033 | 2568 | 359 | 500 | 8 |
| **Apr 23** | 208 | 877 | 44 | 2886 | 773 | 424 | 751 | 16 |
| **Apr 24** | 95 | 1229 | 68 | 2506 | 548 | 251 | 601 | 7 |
| **Apr 25** | 474 | 717 | 22 | 1786 | 641 | 379 | 543 | 12 |
| **Apr 26** | 467 | 749 | 28 | 1976 | 287 | 311 | 434 | 13 |
| **Apr 27** | 199 | 321 | 41 | 1115 | 231 | 208 | 292 | 3 |
| **Apr 28** | 38 | 422 | 28 | 1243 | 402 | 269 | 488 | 6 |
| **Apr 29** | 269 | 516 | 19 | 1197 | 134 | 156 | 268 | 26 |
| **Apr 30** | 308 | 431 | 11 | 1133 | 105 | 128 | 259 | 4 |

Supplementary Table 6. The Omicron cases peaking date、EI and population of each district in Shanghai.

| **District** | **the date of peaking** | **Effective Interval (EI)** | **Population (*10000)** |
| --- | --- | --- | --- |
| **Jingan** | Apr 21, 2022 | 20 | 97.57 |
| **Huangpu** | Apr 20, 2022 | 19 | 66.2 |
| **Pudong** | Apr 13, 2022 | 16 | 568.15 |
| **Hongkou** | Apr 14, 2022 | 13 | 75.75 |
| **Changning** | Apr 12, 2022 | 11 | 69.31 |
| **Putuo** | Apr 11, 2022 | 10 | 123.98 |
| **Xuhui** | Apr 11, 2022 | 10 | 111.31 |
| **Yangpu** | Apr 11, 2022 | 10 | 124.25 |
| **Chongming** | Apr 07, 2022 | 10 | 63.79 |
| **Minhang** | Apr 11, 2022 | 10 | 265.35 |
| **Jinshan** | Apr 06, 2022 | 9 | 82.28 |
| **Baoshan** | Apr 09, 2022 | 8 | 223.52 |
| **Songjiang** | Apr 09, 2022 | 8 | 190.97 |
| **Qingpu** | Apr 09, 2022 | 8 | 127.14 |
| **Jiading** | Apr 08, 2022 | 7 | 183.43 |
| **Fengxian** | Apr 03, 2022 | 6 | 114.09 |

Population information is from the seventh national population census

Supplementary Table 7. Daily subway ridership of each district in Shanghai.

| **District** | **Daily subway ridership** | | | | |
| --- | --- | --- | --- | --- | --- |
|  | **Apr 18, 2015**  **Saturday** | **Apr 19, 2015**  **Sunday** | **Apr 22, 2015**  **Wednesday** | **Apr 23, 2015**  **Thursday** | **Apr 24, 2015**  **Friday** |
| **Pudong** | 1377457 | 1152531 | 1973473 | 1962125 | 2071716 |
| **Xuhui** | 886461 | 734761 | 1211410 | 1191835 | 1264453 |
| **Jingan** | 681097 | 597587 | 941280 | 942166 | 996443 |
| **Huangpu** | 782415 | 627119 | 893713 | 886328 | 953056 |
| **Minhang** | 663753 | 597381 | 831994 | 823760 | 893802 |
| **Changning** | 500679 | 412097 | 739678 | 732458 | 768845 |
| **Putuo** | 436150 | 366340 | 595978 | 593085 | 613605 |
| **Baoshan** | 399426 | 338882 | 520456 | 517868 | 541181 |
| **Hongkou** | 361236 | 300660 | 492646 | 493499 | 513425 |
| **Yangpu** | 323327 | 266286 | 388740 | 386820 | 415962 |
| **Songjiang** | 217269 | 186576 | 250004 | 246818 | 269363 |
| **Jiading** | 203319 | 177323 | 232934 | 232032 | 247363 |
| **Qingpu** | 56896 | 57773 | 106088 | 102790 | 110319 |
| **Fengxian** | 0 | 0 | 0 | 0 | 0 |
| **Jinshan** | 0 | 0 | 0 | 0 | 0 |
| **Chongming** | 0 | 0 | 0 | 0 | 0 |

Subway ridership dataset of Shanghai was shared by Chinese Software Developer Network.

Supplementary Figure 1. The change of time-dependent effective reproductive number in Shanghai’s Omicron wave.

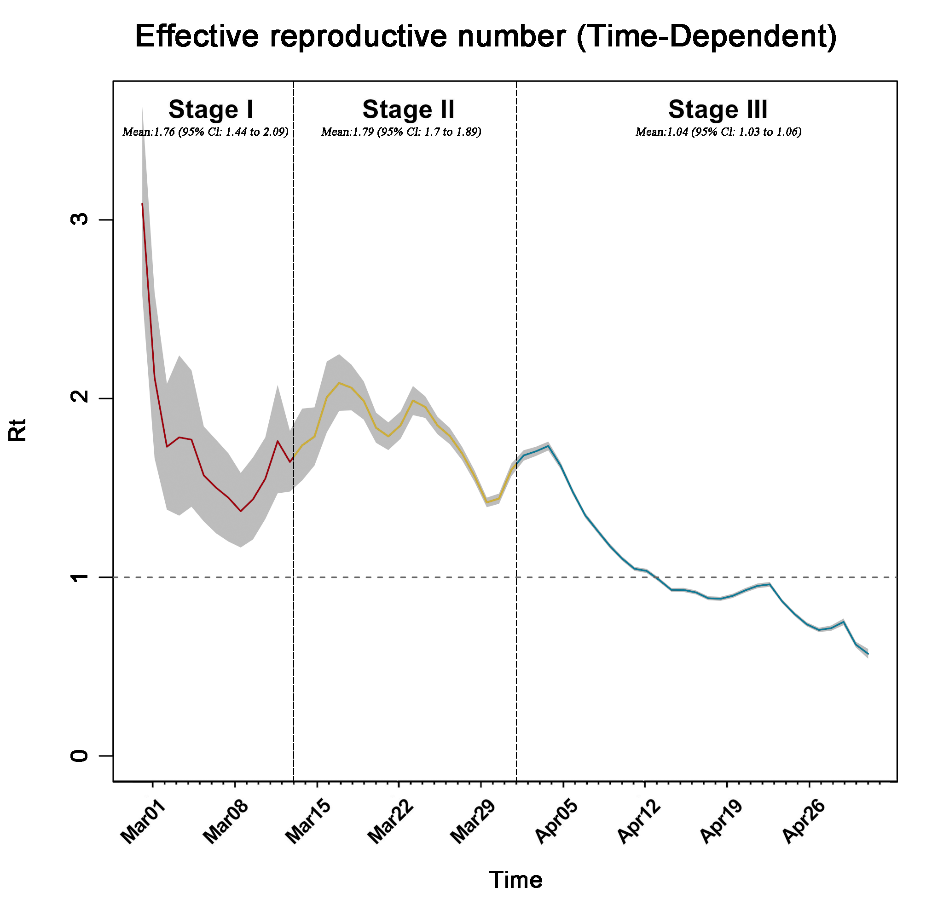

Supplementary Figure 2. Distribution of subway stations whose daily ridership on five days in April, 2015 were used for population mobility calculation in each district.

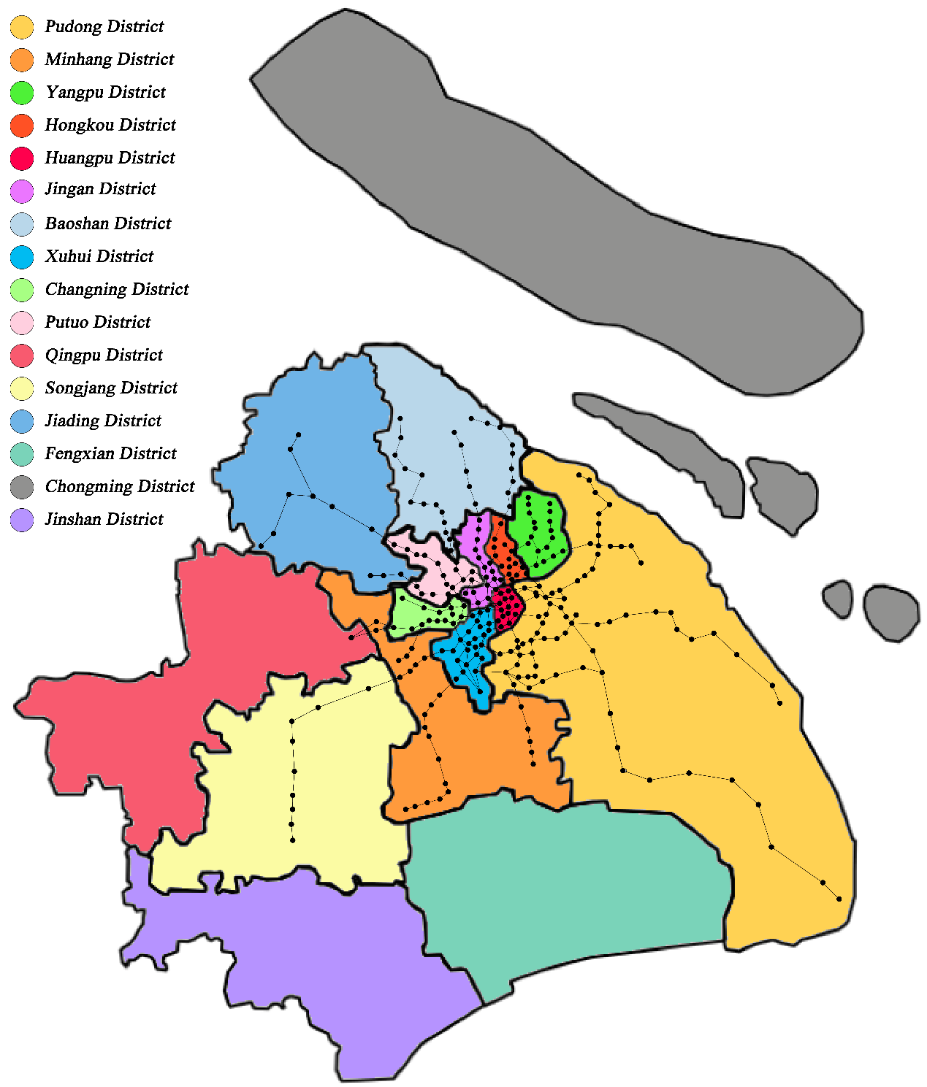
